# Diagnosis Of Satoyoshi Syndrome Using A Neuroblastoma Cell (SH-SY5Y) Lysate As Substrate For Western Blot

**DOI:** 10.1101/2024.11.07.24316705

**Authors:** José María Sevilla Avendaño, Carlos Garrido, Irene Rodríguez Clemente, Julián Solís-García del Pozo, Ulrich Stephani, Ricardo Martínez, Eiji Matsuura, Carlos de Cabo, Valentín Ceña, Javier Solera

**Affiliations:** Research Department, Neuropsychopharmacology Unit, Complejo Hospitalario Universitario de Albacete, Albacete, Spain; Unidad Asociada Neurodeath, Institute of Molecular Nanoscience, INAMOL, University of Castilla-La Mancha, Spain; Department of Internal Medicine, Complejo Hospitalario Universitario de Albacete, Albacete, Spain; Klinik für Klinik für Neonatologie, Kinderpneumologie und Neuropädiatrie (Kinder und Jugendmedizin II, University Hospital of Schleswig Holstein (UKSH), Campus of Kiel, Kiel, Germany; Department of Oral and maxillofacial surgery. Hospital Universitario de Canarias, Santa Cruz de Tenerife, Spain; Graduate School of Medical and Dental Sciences, Kagoshima University, Kagoshima, Japan; Department of Medical Sciences, Faculty of Medicine, Universidad de Castilla—La Mancha, Albacete, Spain

**Keywords:** Satoyoshi syndrome, SH-SY5Y, Western blot, autoimmune, autoantibodies, diagnosis, rare diseases, serum biomarkers

## Abstract

**Objective:** Satoyoshi syndrome is a rare, likely autoimmune disorder that is traditionally diagnosed based on clinical criteria: painful muscle spasms, diarrhea and alopecia. Two previous reports showed a specific immunoreactive band in three patients using Western blot analysis with brain homogenate. The aim of this study was to evaluate the efficacy of SH-SY5Y cell lysate as a potential substitute for brain homogenate in the diagnosis of Satoyoshi syndrome.

**Methods:** Western blot analyses were conducted using brain homogenate, SH-SY5Y cell lysates, and differentiated SH-SY5Y cell lysates. Serum samples were obtained from three patients diagnosed with Satoyoshi syndrome, alongside control samples from thirty blood donors and six patients with other neurological conditions.

**Results:** Sera from patients with Satoyoshi syndrome displayed a consistent three band pattern in the 70–100 kDa range. This pattern was reproducible across all tested substrates (brain homogenate, SH-SY5Y lysate, and differentiated SH-SY5Y lysate), but it was not observed for the sera from the control groups. The immunoreactive bands were more patent when using either SH-SY5Y lysate compared to brain homogenate. No differences were found between the SH-SY5Y lysate and differentiated SH-SY5Y lysate.

**Conclusion:** SH-SY5Y cell lysate could be an alternative to brain homogenate for the immunodiagnostic evaluation of Satoyoshi syndrome. The use of SH-SY5Y cell lysate for Western blot analysis may improve visualization and reproducibility and have a lower cost.

*Key messages:*

- Satoyoshi syndrome is a rare disease currently diagnosed based only on clinical criteria
- A biological marker test for Satoyoshi syndrome could lead to early diagnosis and treatment thus improving the outcomes.
- SH-SY5Y cell lysate may be a better substrate than brain homogenate for Western blot based diagnosis of Satoyoshi syndrome.

## Introduction

Satoyoshi syndrome (OMIM: 600705) [1,2] is a rare, multisystemic disorder characterized by a spectrum of clinical features, including painful intermittent muscle spasms, diarrhea, alopecia, skeletal abnormalities, growth retardation, and endocrinopathies [3, 4, 5, 6]. A delay in diagnosis and the subsequent lack of appropriate treatment can lead to poor outcomes, including severe complications and potentially fatal progression [7, 8]. It is believed that Satoyoshi syndrome is an autoimmune disease. The association of Satoyoshi syndrome with other autoimmune diseases (thyroiditis, myasthenia, systemic lupus, atopic dermatitis, idiopathic thrombocytopenic purpura), the presence of autoantibodies (ANAs, anti-acetylcholine receptor, anti-glutamic acid decarboxylase, among others), and the improvement of symptoms with immune response modifiers are considered evidence supporting an autoimmune origin [3]. The above mentioned serum antibodies are not specific to Satoyoshi syndrome. However, an antibody against brain homogenate has been reported in three previous cases using Western blot that was not found in healthy controls. Endo et al. 2001 described a teenager female patient with Satoyoshi syndrome who carried the anti-AChR autoantibody without presenting symptoms of myasthenia gravis. Analysis of this patient’s serum using Western blot on brain homogenate revealed an 85 kDa immunoreactive band, although the specific antigen remained unidentified [9]. Similarly, Matsuura et al. 2007 reported two female patients (aged 17 and 36) with classic symptoms of Satoyoshi syndrome, whose sera showed a 90-kDa band when tested with brain, stomach, and duodenum homogenates using Western blot [10].

The aim of this study was to evaluate neuroblastoma SH-SY5Y cell lysate as a substitute for brain homogenate for Satoyoshi syndrome diagnosis by Western blot. The use of SH-SY5Y cell lysate may improve visualization and reproducibility and have a lower cost.

## Material and Methods

### Patients’ samples

The serum from three patients with Satoyoshi syndrome previously reported were included in this study [11, 12, 13]. A brief summary of the three cases follows:

#### Case 1 [11]

A woman in her 20’s from Spain developing alopecia in her late childhood. Two years later, she developed progressive painful muscle spasms, followed by diarrhea with carbohydrate malabsorption, iron-deficiency anemia, weight loss, skeletal abnormalities and amenorrhea. A definitive Satoyoshi syndrome diagnosis was not reached until the end of her teenage. Treatment with prednisone (30 mg/day) and methotrexate (7.5 mg/week) was initiated with dramatic improvement of muscle spasms and resolution of diarrhea, gaining 20 kg and recovered menstruation.

#### Case 2 [12]

A teenager woman from Spain with a history of asthma and allergic rhinitis that presented with universal hair loss in her early childhood. In her middle childhood, she exhibited painful, intermittent muscle spasms, significant growth retardation, diarrhea, and abdominal pain. Her muscle spasms increased with physical activity and were localized to the acral areas of her limbs, wherein they typically lasted several minutes and were self-limited. The patient was diagnosed with Satoyoshi syndrome. Carbamazepine and otilonium bromide treatment led to the disappearance of her muscle spasms and diarrhea.

#### Case 3 [13]

A woman in her 30’s from Germany. As an adolescent, she started muscle spasms, jaw cramps being the most uncomfortable. She was treated with carbamazepine, prednisolone, and sex-steroids. After introduction of low-doses of methotrexate to the therapy the patient recovered from muscle spasms, alopecia and diarrhea. At the end of her teenage, medication of MTX and corticosteroids was tapered off without relapse. Initiation of sex-steroid treatment resulted in pubertal development, regular menstrual cycles. Being married nowadays she experienced three pregnancies with spontaneous abortions.

### Control groups

Two control groups were included: one consisting of 30 blood donors were provided by the Albacete General University Hospital Blood bank and a second group with patients with other neurological pathologies. Briefly, a female in her 50’s with radiculopathy, a male in his 40’s with acute disseminated encephalomyelitis, a female in her 50’s with secondary progressive multiple sclerosis, a male in his 70’s with Parkinson’s disease, a female in her 30’s with encephalitis caused to human herpes virus type 6, and a male in his 50’s with demyelinating polyneuropathy were included in the study. Samples from these patients were provided by the Albacete General University Hospital Biobank.

The present study was compliant with the Declaration of Helsinki and ethical approval was obtained prior to data and sample collection by the local Medicine Research Ethics Committee of the Albacete General University Hospital (Acta 04/2016 and Acta 10/2021). All participants provided written informed consent for this study.

### Laboratory methods

For the identification of autoantibodies in patient serum, we used whole brain homogenate (Biochain, Newark, CA, USA) and lysates from SH-SY5Y (ATCC, Manassas, VA, USA) cells and differentiated SH-SY5Y cells. The brain homogenate was reconstituted according to the manufacturer’s instructions.

### Preparation of the SH-SY5Y cell lysates

SH-SY5Y cells were cultured in Dulbecco’s Modified Eagle Medium (DMEM F-12) supplemented with 2 mM L-glutamine (Gibco, Grand Island, NY, USA), 100 U/mL penicillin and 100 U/mL streptomycin (penicillin-streptomycin; Gibco, Grand Island, NY, USA), and 10% (v/v) heat-inactivated fetal bovine serum (FBS; Gibco, Gaithersburg, MD, USA). Cultures were grown in a humidified incubator at 37°C under a 5% CO2 atmosphere. To induce differentiation, SH-SY5Y cells were treated with 0.1% retinoic acid (Sigma-Aldrich, Saint Louis, MO, USA) for 5 days, with the medium changed every 2 days. The culture medium of SH-SY5Y cells was removed, and the cells were washed with ice-cold phosphate-buffered saline (PBS, 0.1 M phosphate, 0.15 M sodium chloride, pH 7.2) and lysed for 5 minutes in 300 μL of ice-cold lysis buffer (Pierce IP, Thermo Fisher Scientific, Rockford, IL, USA). The cell lysates were centrifuged at 13,000 g for 10 minutes. The supernatants were kept for Western blot analysis and the pellets discarded.

### Western blot

Protein concentrations were determined spectrophotometrically using the Micro BCA Protein Reagent Kit (Thermo Fisher Scientific, Rockford, IL, USA). A total of 25 µg of protein from brain homogenate, SH-SY5Y lysate, and differentiated SH-SY5Y lysate were separated by electrophoresis on 8% SDS-PAGE gels, 30 minutes at 60V and 1.5 - 2 hours at 100V. Following electrophoresis, proteins were transferred to nitrocellulose membranes (Immobilon; Millipore Corporation, Billerica, MA, USA) previously activated with transfer buffer; 3 gr. of Base Trizma (Sigma-Aldrich, Saint Louis, MO, USA), 14.4 gr. of glycine (Sigma-Aldrich, Saint Louis, MO, USA), 200 mL of methanol and 800 mL of milliQ water; in a Trans-Blot® SD Semi-Dry Transfer Cell (Bio-Rad, Hercules, CA, USA) at a constant amperage of 20 mA during 1.5 hours. Nonspecific protein binding was blocked using a blocking buffer, 10% w/v skimmed milk and 0.1% Tween 20 (Sigma-Aldrich, Saint Louis, MO, USA) in PBS for 1 hour at room temperature (RT). The membranes were incubated overnight at 4 °C with patient serum samples or control serum samples, diluted 1:50 in a blocking buffer. After washing with PBS containing 0.1% v/v Tween 20 x5 times for 5 minutes, the membranes were incubated with an anti-human IgG secondary antibody (Anti-Human IgG (Fc specific)−Peroxidase antibody produced in goat; Sigma-Aldrich, Saint Louis, MO, USA) diluted 1:1000 in blocking buffer at RT and washed again with PBS containing 0.1% v/v Tween 20 x5 times for 5 minutes. Antibodies were detected using an enhanced chemiluminescence detection kit (GE Healthcare, Little Chalfont, Buckinghamshire, UK). The images were acquired using the transilluminator (Luminescent Image Analyzer LAS-4000 mini, Fujifilm, Tokio, Japan) with the image capture software (LAS-4000 Image Reader, Fujifilm, Tokio, Japan).

## Results

In a first experiment, we assessed the autoimmune immunoreactivity of the serum from the Satoyoshi patients in brain homogenate and SH-SY5Y cell lysate. We observed a pattern of three bands between 70 and 100 kDa in both brain homogenate and SH-SY5Y lysate samples. A better intensity and definition of the bands was observed for the SH-SY5Y lysate (Fig 1a). An additional western blot was performed to compare SH-SY5Y lysate and differentiated SH-SY5Y lysate. The same band pattern was found for both lysates, but no difference in the band intensity and definition (Fig 1b).

**Figure 1.**
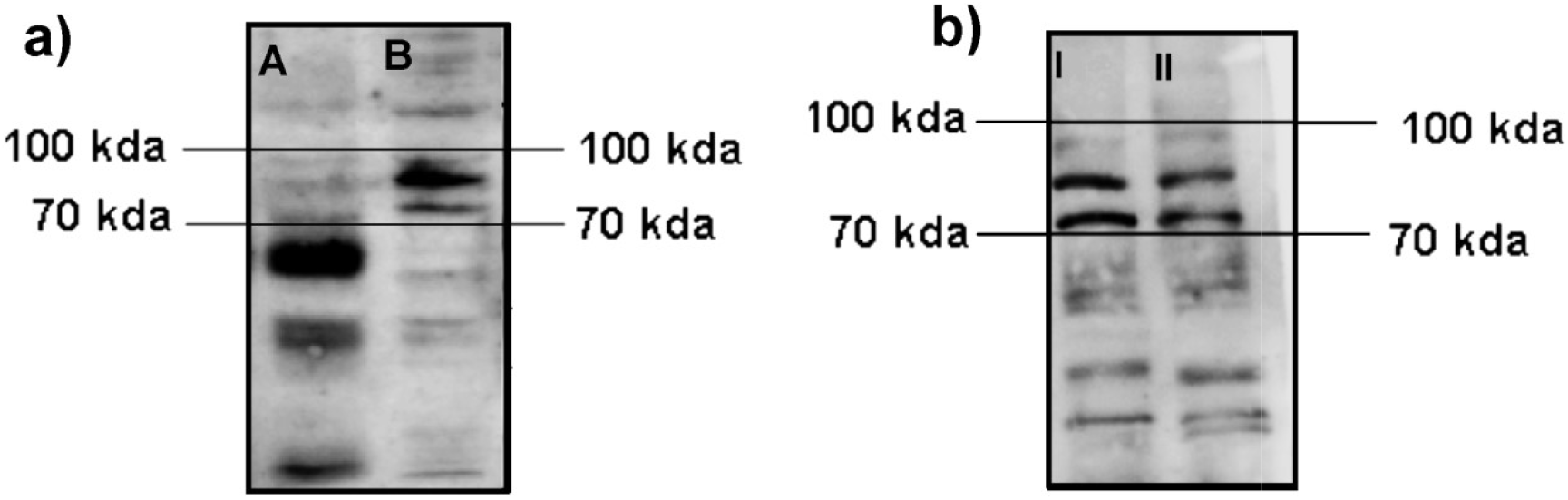
Comparisons of Western blot immunoreactivity results using brain homogenate vs SH-SY5Y lysate and SH-SY5Y lysate vs differentiated SH-SY5Y lysate. A) after incubation with serum from the Satoyoshi patient case 1 for brain homogenate (A) and SH-SY5Y cell lysate (B), a common band pattern between 70 kDa - 100 kDa was found. b) Comparison between the SH-SY5Y cell lysate (I) and the SH-SY5Y differentiated cell lysate (II) revealing the same band pattern between 70 kDda - 100 kDa. A longer running time (2 hours vs 1,5 hour) was employed for this blot to allow a better separation of the bands.

In a second experiment using SH-SY5Y lysate samples, we observed a common band pattern between 70 kDa and 100 kDa for the serum samples of the three Satoyoshi syndrome patients (Fig. 2, E=case 1, F= case 2, G= case 3). This band pattern was absent for blood donors(Fig. 2, A-D) and for patients with other neurological conditions (Fig. 2, H-M), suggesting that this band pattern is specific to Satoyoshi syndrome.

**Figure 2:**
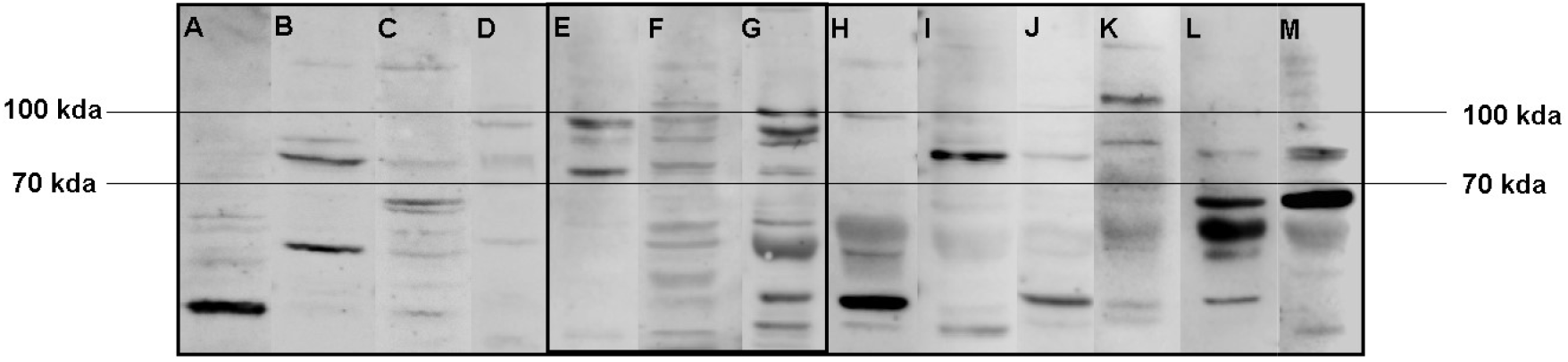
Comparisons of Western blot immunoreactivity band pattern between patients with Satoyoshi syndrome and control groups using SH-SY5Y lysate. After incubation with serum from blood donors (A-D), serum from Satoyoshi patients (E=case 1, F= case 2, G= case 3) and serum from patients with other neurological conditions (H-M) for SH-SY5Y cell lysate, a specific band pattern between 70 kDa - 100 kDa was found for the Satoyoshi patients. This pattern was not observed for either control group.

## Discussion

We have been able to demonstrate that SH-SY5Y cell lysate can serve as a substitute for brain homogenate in the Western blot diagnosis of Satoyoshi syndrome. A characteristic band pattern in our three patients with Satoyoshi syndrome was identified, which was more patent using the SH-SY5Y cell lysate than using the brain homogenate. No differences were found between the results from SH-SY5Y cell and differentiated SH-SY5Y cell lysates. This immunoreactive band pattern was absent for serum from blood donors or patients with other neurological conditions. The band pattern found in our study, between the 70-100 kDa, is within the range of the bands described by Endo et al. (85kDa) and Matsuura et al. (90 kDa) employing a similar Western blot technique [7,8]

The serum samples from this study were obtained at least 5 years after the onset of the disease, when the patients had already received treatment and no longer showed symptoms. The persistence of these characteristic autoantibodies after treatment may allow a retrospective diagnosis. Satoyoshi syndrome is a very rare disease, its symptomatology takes years to fully develop and the appearance of the different symptoms is heterogeneous across patients. Up to now, diagnosis is based on a typical clinical pattern. This western blot test could provide an early diagnosis, e.g. when patients only present with alopecia or muscle spasms. However, samples from more patients are needed to confirm the validity of the test. Additionally, efforts should be made to identify the autoantibodies responsible for this immunoreactivity as well as the target protein or proteins.

In summary SH-SY5Y lysate is a suitable substitute for human brain homogenate for the diagnosis of Satoyoshi syndrome using the Western technique. Its advantages include improved band visualization, better reproducibility, and lower cost. Additional studies involving more Satoyoshi syndrome patients are needed to validate these results.

## Data Availability

All data produced in the present study are available upon reasonable request to the authors

## Acknowledgements

We thank all patients and blood donors for agreeing to take part in this research. Many thanks to the Biobank of the Albacete General University Hospital, integrated into the National Biobank Network of the Carlos III Institute of Biomedical Research, for kindly supplying the samples from patients with other neurological conditions for control. Our thanks also go to the Albacete General University Hospital Blood bank for collecting the samples from the blood donors.

## Author Approval

All authors have seen and approved the manuscript.

## Conflict of interest statement

The authors have declared no conflicts of interest.

### Data availability statement

The raw data supporting the conclusions of this article will be made available by the authors, without undue reservation.

